# Using flexible carbon fiber insoles for 1^st^ metatarsophalangeal arthritis lead to pain reduction and high compliance rate: A Randomized Controlled Trial

**DOI:** 10.1101/2021.04.05.21253422

**Authors:** Pongpanot Sornsakrin, Rohan Bhimani, Michael Drew Vrolyk, Bart Lubberts, Daniel Guss, Christopher W DiGiovanni, Gregory Waryasz

## Abstract

**Introduction:** Shoe modification and orthotics play an important role in non-operative management for 1^st^ metatarsophalangeal (MTP) arthritis. However, compliance can be low due to pain and discomfort. We hypothesized that patients who wear a flexible carbon fiber insole for 1^st^ metatarsophalangeal arthritis will report reduced pain, and higher physical function and compliance rate when compared with a rigid Morton’s extension insole.

**Methods:** Four males and nine females (mean age of 56 years; range 35-79) diagnosed with 1^st^ metatarsophalangeal arthritis were included in this randomized controlled trial. Participants randomly received either bilateral flexible carbon fiber insoles (VKTRY®) (n=7) or unilateral Morton’s extension insole (n=6). Outcome measures included patient compliance, comfort rate, and Patient-Reported Outcomes Measurement Information System (PROMIS) questionnaires Global Health, pain interference, v1.0 pain intensity, v2.0 Physical Function, and v1.0 Depression. Participants filled out questionnaires at baseline and at two, six, and twelve weeks follow-up.

**Results:** Compared to Morton’s extension insole, use of flexible carbon fiber insole led to reduction of pain interference score at 6 and 12 weeks (median ⍰ −9.5 vs 0.0 p=0.015; and median ⍰ −15.1 vs −2.3 p=0.015, respectively), as well as reduction of pain intensity score at 6 and 12 weeks (median ⍰ −11.9 vs −2.3 p=0.018; and median ⍰ −11.9 vs −2.3 p=0.010, respectively). The compliance rate in flexible carbon fiber insole group was 100% at 2, 6 and 12 weeks, compared to 83%, 83% and 50% in the Morton group. In addition, patients wearing carbon fiber insoles experienced higher comfort levels (p-values ranging from <0.001 to p=0.007). There were no differences between the comparison groups at 2, 6, and 12 weeks in terms of the global health, physical function, and depression scores.

**Conclusion:** Patients diagnosed with 1^st^ metatarsophalangeal arthritis may benefit from wearing flexible carbon fiber insoles, compared to commonly used rigid orthotic insoles, such as the Morton’s extension. This insert can be used safely for nonoperative treatment of hallux rigidus and is another option for nonoperative treatment.

**LEVEL OF EVIDENCE:** Level I, randomized clinical trial.

## Introduction

Hallux rigidus is a degenerative condition of the first metatarsalphalangeal (1^st^ MTP) joint, affecting 2.5% of people over the age of 50.(1) The prevalence of symptomatic radiographic osteoarthritis of the 1^st^ MTP joint (radiographic changes with associated clinical symptoms) in people over the age of 50 years is 7.8%, with a higher prevalence observed in women, older people, and those from lower socio-economic classes, while 72% of people with symptomatic 1^st^ MTP osteoarthritis reported disabling symptoms.(2) Non-operative management includes the use of NSAIDS, intra-articular injections, shoe modification, activity modifications, and physical therapy.^1^ Shoe modification and orthotics reduce pain by modifying the biomechanics of the first MTP joint.^1^ However, compliance of patients wearing an orthotic device (Morton extension) can be low due to pain and discomfort.^2^ The traditional orthotic aims to decrease the range of motion of the joint while walking. Flexible carbon fiber insoles are shock absorbing and were initially designed to increase ground force leading to a harder push off for faster running or higher jumping. They have been touted to possibly improve performance but have not been studied for specific conditions of the foot and ankle to date.

The aim of this study was to compare a flexible carbon fiber insole with a rigid Morton extension insole in a single-blinded randomized controlled trial. We hypothesized that patients who wear a flexible carbon fiber insole for 1^st^ metatarsophalangeal arthritis report reduced pain, and higher physical function and compliance rate when compared with a rigid Morton’s extension insole.

## Materials & Methods

### Study Design and Setting

This randomized controlled trial was approved by our Institutional Review Board under protocol #2018P002864. Patients were enrolled at two hospitals between April 2019 and February 2020. Signed informed consent was obtained from all subjects.

### Study participants

Inclusion criteria were adult patients equal or older than 18 years of age diagnosed with unilateral 1^st^ MTP joint arthritis based on clinical exam and radiographic imaging, and patients who were competent and able to consent on their behalf. We excluded patients who had history of 1^st^ MTP joint injection, prior surgery to the foot, prior use of orthotic, or need for surgical treatment. When participants were enrolled, the researcher delivered the orthosis directly to the participant within one week at no cost.

### Randomization

Once informed consent was obtained, patients were randomized into one of two groups; receiving either bilateral flexible carbon fiber insoles (VKTRY®, Milford, CT, USA) or unilateral rigid Morton’s extension insole (Dr. Jill’s Foot Pads Inc., Deerfield Beach, FL, USA) for a total period of 12 weeks. Patients were not told that the rigid Morton extension insoles is the control group and that the flexible carbon fiber insoles group was the study arm. A block randomization was performed by alternating computer-generated blocks with a size of four. Patients were asked to wear the insole during daily activities as tolerated.

### Explanatory variables

The following patient and disease data were obtained from each study participant: age, sex, weight, height, ethnicity, range of motion of the 1^st^ metatarsophalangeal joint, and comorbidities at time of enrollment. All patients had plain radiographs confirming the diagnosis as well. There was no missing data.

### Outcome Measures

The primary outcome measure was the Patient Reported Outcomes Measured Institute System (PROMIS) pain score, which included the pain interference and pain intensity domain. The PROMIS(r) Pain Interference instrument measures the self-reported consequences of pain on relevant aspects of a person’s life and may include the extent to which pain hinders engagement with social, cognitive, emotional, physical, and recreational activities. Pain Interference also incorporates items probing sleep and enjoyment in life, though the item bank only contains one sleep item. The Pain Interference short form is universal rather than disease-specific. The Pain Interference items utilize a 7–day recall period (items include the phrase “the past 7 days”).(3-5) On the other hand, the Pain Intensity short form (3a) determines psychometric properties and clinical input. The Pain Intensity short form is universal rather than disease-specific. The first two items in the Pain Intensity item bank assess pain intensity utilizing a 7–day recall period (items include the phrase “the past 7 days”) while the last item asks patient to rate their pain intensity “right now.”(4, 5) The PROMIS v2.0 is drawn from a bank of 17 items that focus exclusively on lower extremity function.10 Each participant answers 4–12 items; the CAT discontinues when a preset level of precision is achieved (SE # 3.0) or 12 items are completed, whichever happens first. The PROMIS v1.0 Physical Function Short Form 8a (PFSF8a) includes 8 items selected from the PROMIS Physical Function item bank.9 Items reflect mobility, upper extremity function (eg, carrying a bag of groceries), and full-body activities (eg, run errands and shop). For both PROMIS measures, the mean T-score in the United States general population is 50 with a SD of 10. Higher scores reflect better physical function. Higher scores reflect better physical function. Mobility computer adaptive tests (CAT) scores range from 18 to 60 and the PFSF8a scores range from 20 to 60. The patient responded by using five-category response scales (without any difficulty, with a little difficulty, with some difficulty, with much difficulty, unable to do).(6) The PROMIS Global health was assessed to understand how patient’s symptoms affected physical function, pain, fatigue, emotional distress, social health. The PROMIS global health consists of 10 items with 9 items using five-category response scales, and one item (rating of pain on average) using a response scale of 0–10 that is recoded to five categories (0 = 1; 1-3 = 2; 4-6 = 3; 7-9 = 4; 10 = 5).(7) The PROMIS Bank v1.0 depression; these items in the questionnaire focus on the negative moods, emotions, self-image, social cognition and cognitive deficits. A 5-point verbal scale was used to measure patients experience related to their symptoms (“I felt depressed: never, rarely, sometimes, often, always”).(6) Furthermore, patient compliance whether the patient used the orthosis – or not. This was a dichotomous choice that the patient would response “Yes” or “No” which referred to whether they used the orthosis at the day they completed the questionnaire. Lastly, the comfort rate was measured with a 0-100 Likert scale, where a score of 0 indicated “least comfort” and a score of 100 reflected “best comfort”. All outcome measures were self-reported by the patients at baseline and at 2, 6, and 12 weeks during regular office visits. There was no missing data.

### Sample size and statistical analysis

To test the null hypothesis that there was no difference in PROMIS PF scores between the comparison groups, we used a t-test. Hung et al. found that in foot and ankle patients the minimal clinically important difference (MCID) for the PROMIS PF ranged from 7.9 to 13.2 at 3-month follow-up and beyond, with a standard deviation (SD) ranging from 4.5 to 4.7.(8) In order to achieve 80% statistical power for detecting a difference of 10.6 (+/-4.6 SD) points of the MCID in the PROMIS pain intensity score between patients using the Carbon fiber insole and patients using the Morton’s extension insole with an overall two-tailed Type-1 rate of 5%, and accounting for a 30% lost to follow up, we would need 14 patients total. The sample size calculation was performed using G*Power Version 3.1.9.2.

The baseline characteristics, age, sex, hallux range of motion, and BMI were obtained and summarized using frequency for categorical variables and mean ± standard deviation (SD) for continuous data. A Mann-Whitney U test was used to compare the increase or decrease of each outcome variable over time (Delta (⍰); baseline score – follow-up score) between the insole groups. This test was applied to test the primary hypothesis that patients who used the flexible carbon fiber insole would have higher improvement of the PROMIS PF score than patients who used Morton’s extension insole. A paired t-test (a parametric test used when comparing two dependent samples) was applied to test the secondary hypothesis that the compliance and comfort rate would be higher in patients wearing the flexible carbon fiber insoles. A p-value < 0.05 was considered as statistically significant. Data analyses were performed using Stata 14.0 (StataCorp LP, College Station, TX)

## Results

Thirteen adult-patients, four male and nine female patients were included (Figure 1, Consort diagram). The mean age of the patients was 56 years (range 35-79). One patient in the Morton’s extension insole group was excluded from the analysis due to operative treatment for persistent pain of the hallux at two weeks follow-up. There were no differences between the flexible carbon fiber insole (n=7) and Morton’s extension insole group (n=6) in terms of patient’s age, gender, hallux range of motion, radiographic severity of hallux rigidus (based on Hattrup and Johnson classification) and BMI at baseline (Table 1).

**Figure 1.**
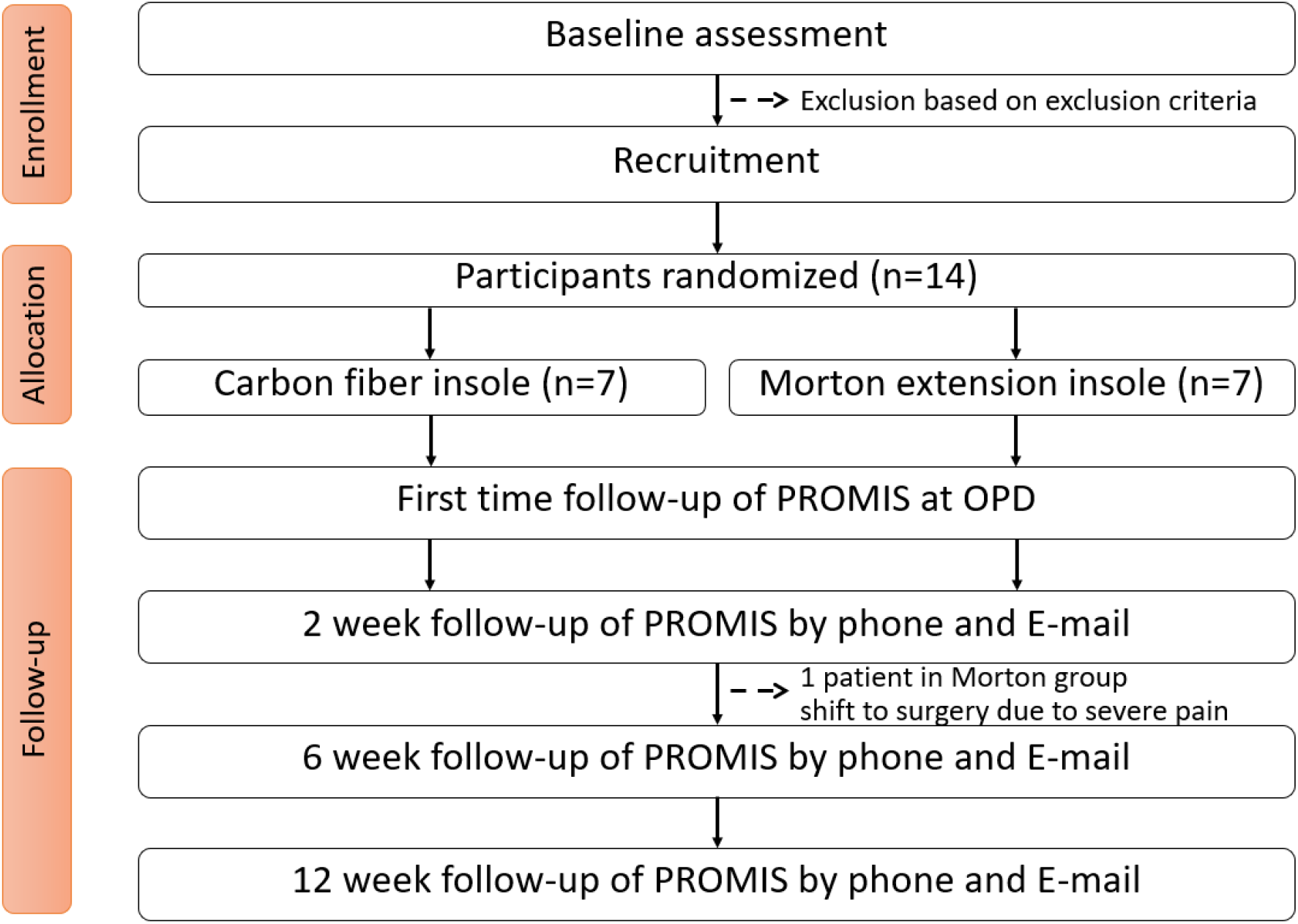
The CONSORT diagram of patients enrolled and randomized in either the Morton and Carbon fiber insole groups.

**Table1.** The demographic data of the participants shown no difference between the rigid morton- and flexible carbon fiber group.

| | | Morton Insoles<br>(mean ± SD) | Carbon Fiber Insoles<br>(mean ± SD) | P-value |
| --- | --- | --- | --- | --- |
| Age (years) | | 57.67 ± 14.19 | 53.29 ± 7.80 | 0.496 |
| Gender | Male | 2 | 2 | 0.853 |
|  | Female | 4 | 5 |  |
| Grade of arthritis on<br>x-ray | Grade 1 | 3 | 3 | 0.939 |
|  | Grade 2 | 2 | 3 |  |
|  | Grade 3 | 1 | 1 |  |
| Range of motion (degree) | | 25.83 ± 6.76 | 24.29 ± 3.17 | 0.842 |

**Table 2.**
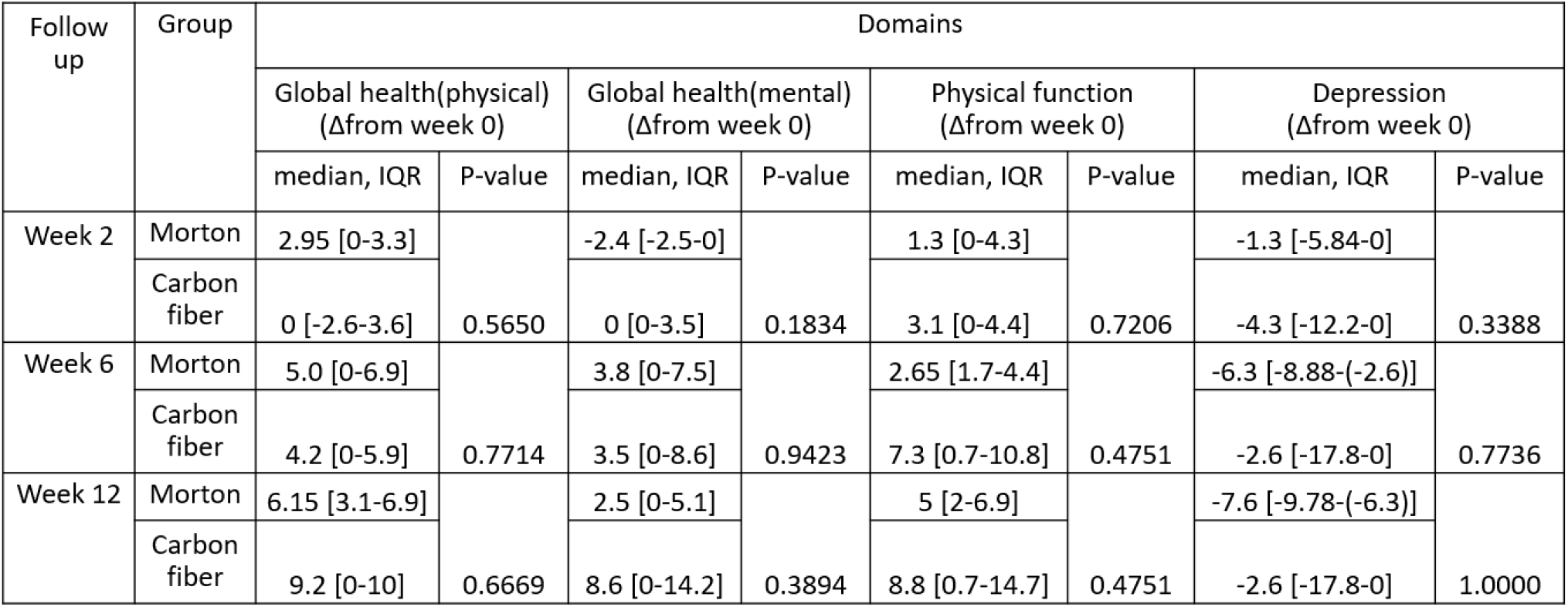
The PROMIS Global physical health, Global mental health, Physical function and Depression scores at 2, 6, and 12 weeks follow-up (⍰; baseline score – follow-up score) among the rigid morton- and flexible carbon fiber insole groups.

Compared to Morton’s extension insole, use of the flexible carbon fiber insole led to a reduction of the pain interference score at 6 and 12 weeks (median ⍰ −9.5 vs. 0.0 p=0.015; and median ⍰ −15.1 vs. −2.3 p=0.015, respectively), as well as a reduction of the pain intensity score at 6 and 12 weeks (median ⍰ −11.9 vs. −2.3 p=0.018; and median ⍰ −11.9 vs. −2.3 p=0.010, respectively (Figure 2). All patients in the flexible carbon fiber insole group continued to use their orthosis until the end of 12 weeks, which was higher than the Morton insoles (83.3%, 83.3% and 50% at 2, 6, and 12 weeks). Patients experienced higher comfort levels (p<0.001, p=0.007, p<0.001, respectively (Figure 3). There were no differences between the insole groups at 2, 6, and 12 weeks in terms of the global health, physical function, and depression scores.

**Figure 2.**
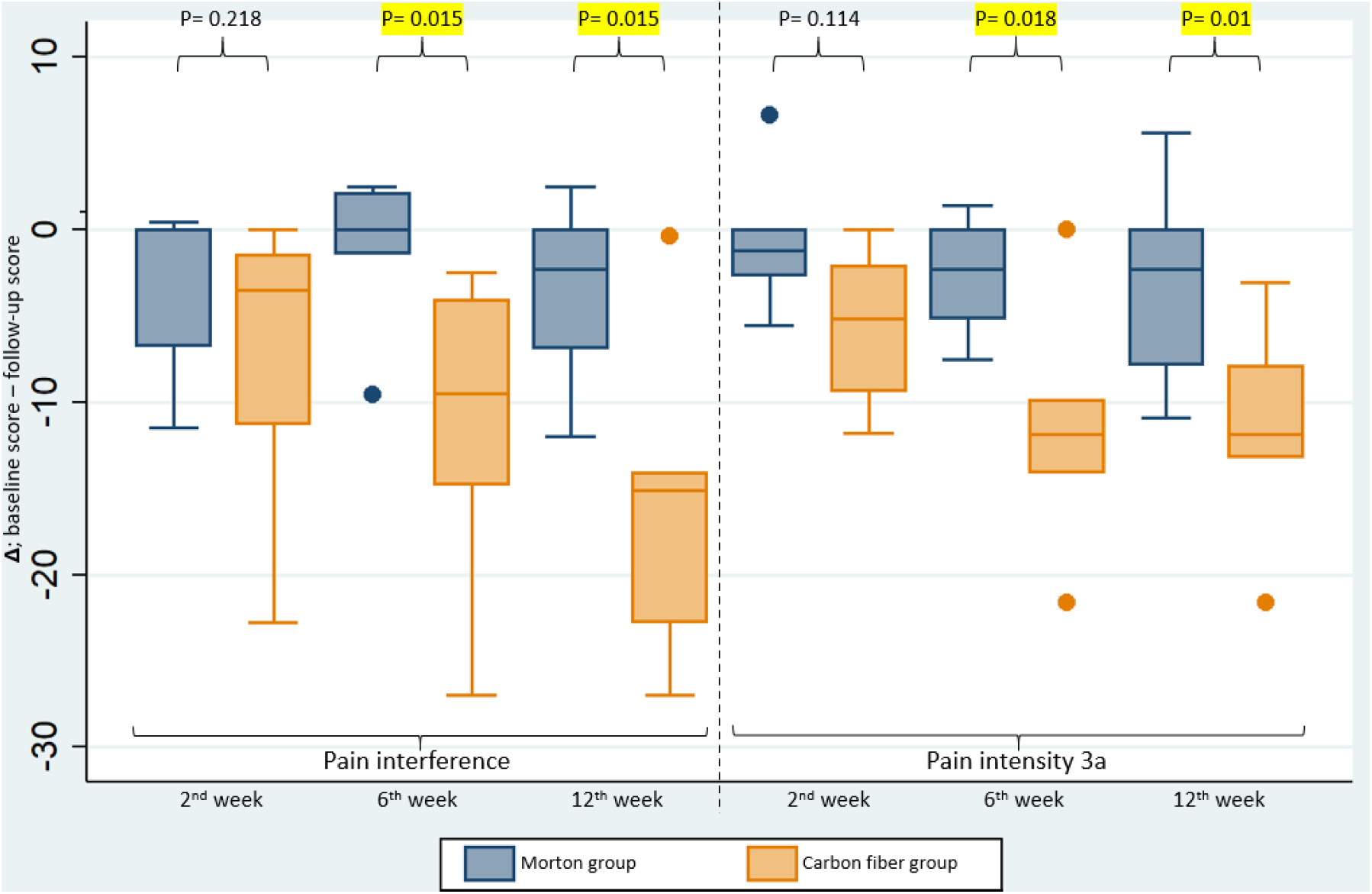
The median increase or decrease of the PROMIS Pain interference and Pain Intensity scores at 2, 6, and 12 weeks follow-up (⍰; baseline score – follow-up score) among the rigid morton and flexible carbon fiber insole groups.

**Figure 3.**
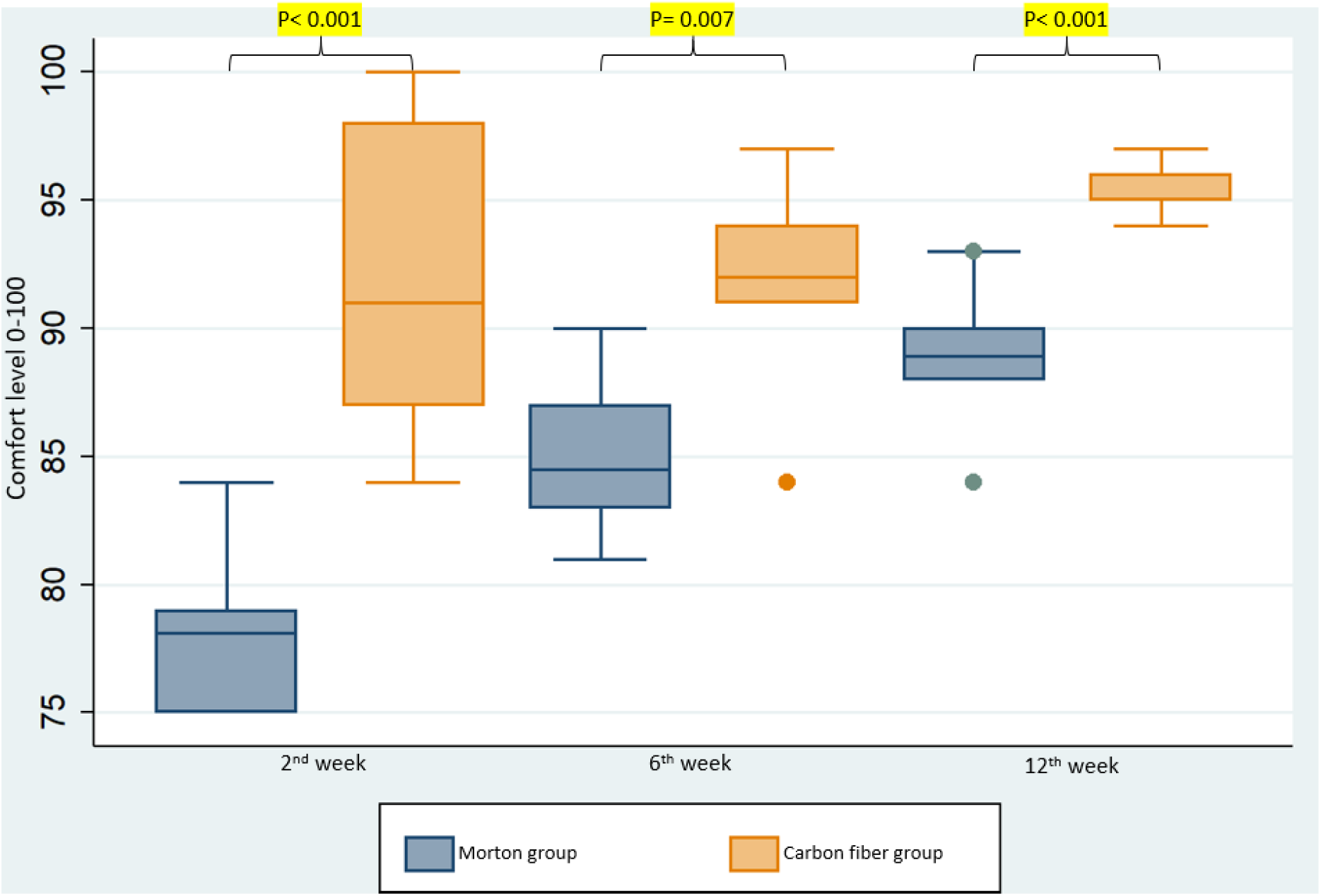
The comfort levels between using of rigid morton- and flexible carbon fiber insoles in the same week of follow-up.

## Discussion

Non-operative treatment using shoe modification and orthotics is common practice for management of the 1st MTP joint arthritis.(1) However, due to pain and discomfort, the compliance rate of patients using these modifications is relatively low.(2) This can cause a reduction of patient foot-specific and general health-related quality of life.(9) Our data suggest that patients who wear flexible carbon fiber insoles have reduced pain and have higher compliance rate than patients who use rigid Morton extension orthosis for 1st MTP joint arthritis.

The treatment goals for first MTP joint osteoarthritis are to reduce pain and stiffness, as well as to prevent further degeneration of the joint.(10) Pain is the main reason why patients with Hallux rigidus look for treatment and is strongly related to functionality. In the present study, patients using the flexible carbon fiber insoles for 1^st^ MTP arthritis over a 12 week period showed a higher reduction of PROMIS pain scores (pain interference and pain intensity) compared to patients wearing Morton’s extensions. In both the groups pain was reduced, with carbon fiber insoles group improved to a greater extent demonstrating clinical effectiveness of the foot orthosis. The pain improvement in the flexible carbon fiber insoles group could be due to the high tensile strength and spring-like hinge action that snaps back after being bent. In both the groups, patients took at least 6 weeks to get accustomed to the insoles which resulted in improved PROMIS scores (Figure 2). However, the comfort level of the Carbon fiber insole was significantly higher than the Morton’s extension insole at 2^nd^, 6^th^, 12^th^ week, respectively (Figure 3).

Patient reported outcomes are increasingly incorporated into clinical practice and provides information about the perspectives of patients and provides a mechanism for evaluating treatment effectiveness that is meaningful to patients. Minimal clinically important difference (MCID) scores are useful to help clinicians and patients gain perspective on relative levels of meaningful change. In a previous study, Hung et al (8) reported that the minimal clinically important difference (MCID) for Pain Interference to be 4.1 to 4.3 using the one-half standard deviation method. Our results showed that the flexible carbon fiber insoles had reached that MCID threshold with a pain interference score of 15.1 suggesting that the carbon fiber insoles to be superior to Morton’s extension. Moreover, in both the groups, significant improvement in physical function was seen compared to baseline, despite no statistical significance between the scores. For physical function, Hung et al (8) reported the MCID to be 4.5 to 4.7 using the one-half standard deviation method and both the carbon fiber insoles (8.8) and Morton’s extension (5.0) cohorts in this study reached that MCID threshold. This suggests that the flexible carbon fiber insoles may provide equivalent or better relief compared to the rigid Morton’s orthosis.

Noncompliance can have significant social and financial impact on the patients. Previous clinical studies have reported excellent outcomes in high compliance patients with 1^st^ MTP joint arthritis using conservative treatment with orthosis. However, compliance of wearing orthosis is reported to be low due to discomfort. (11-13) Poor compliance of orthosis use can lead to a high financial loss for society and a waste of therapeutic effort.(14) Even though Morton’s extension insole can relieve pain at the 1^st^metatarsal joint due to arthritis, the rigid material may interfere with the normal gait and lead to unfavorable everyday use of the orthosis. In contrast, carbon fiber insoles that have a semi-rigid property, may absorb some of the ground reaction forces during gait. This may result in increased comfort and higher compliance than the currently used orthotic device. We opted not to control for shoe wear in the study and patients were allowed to move the inserts into different shoes. Patients were also allowed to use oral and topical pain relivers as needed for pain control.

This study has a few limitations that should be considered. First, the relatively small sample size prohibit us from doing subgroup analyses within the study cohort. For example, it would be of interest to assess differences in PROMIS scores between males and females or patients with various BMI scores. Nevertheless, the sample size was adequate as this was based on a clinically significant PROMIS PF score, which is one of the most commonly assessed PROMIS outcome measures following treatment.(4-6, 15) Notably, although the PROMIS PF score was increased in the flexible carbon fiber orthosis group, compared to the Morton group, this increase may not be clinically relevant, due to the relatively small sample size. Another limitation of the study is a potential bias by use of unilateral vs. bilateral orthosis. The patients in Morton’s extension orthosis group received one orthosis for the affected foot in contrast to those in the carbon fiber insoles group who received orthosis for both feet. However, it mimics how a patient would buy the insole, because the flexible carbon fiber insoles (VKTRY®) is sold as a pair, whereas the rigid Mortons extension is sold as a single side. Finally, patients could not be blinded to the orthosis group they were assigned to. In an attempt to reduce potential placebo effects, all patients were instructed that both the interventions had been shown to improve foot pain and that it was not known if one intervention was more effective than another. Patients were blinded as well to what the study arm was.

## Conclusion

Compared to a rigid Morton’s extension insole for non-operative treatment of 1st MTP arthritis, patients may benefit using flexible carbon fiber insoles (VKTRY®) instead. Using flexible carbon fiber insoles showed a significant reduction of pain scores over time and a higher compliance rate compared to rigid Morton’s extension insole. The advantage of the carbon fiber insoles is likely related to the elasticity of the insole, which may absorb ground reaction forces during gait. Both orthotics did show clinical benefit over baseline PROMIS scores. The flexible carbon fiber is another viable option for nonoperative management of 1^st^ MTP arthritis.

## Data Availability

All data generated or analysed during this study are included in this published article

## Conflict of Interest

This research received non-financial research support from VKTRY.inc, including the flexible carbon fiber insoles and the rigid Morton’s extension insole.

## Notes

### Competing Interest Statement

Dr. Waryasz reports non-financial support from VKTRY, Inc., during the conduct of the study; non-financial support from Arthrex, Inc, outside the submitted work; and American Orthopaedic Foot and Ankle Society: Board or committee member. Dr. DiGiovanni reports non-financial support from Arthrex, Inc, non-financial support from Butterfly Network, non-financial support from OrthoScan, Inc, outside the submitted work; and American Orthopaedic Foot and Ankle Society: Board or committee member, Foot and Ankle International: Editorial or governing board, Foot and Ankle Orthopaedics: Editorial or governing board, Techniques in Orthopaedics: Editorial or governing board. Dr. Guss reports non-financial support Arthrex, Inc, non-financial support from Butterfly Network, non-financial support from Orthoscan, other from AOFAS/DJO Traveling Fellowship, other from Wright Medical Technology, Inc., outside the submitted work; and American Orthopaedic Foot and Ankle Society: Board or committee member. Dr. Lubberts reports non-financial support from Arthrex, Inc, non-financial support from Butterfly Network, outside the submitted work; and Orthopaedic Research Society: Board or committee member. The other authors declare that they have no conflict of interest.

### Clinical Trial

NCT04833608

### Author Declarations

The study was approved by Mass General Brigham IRB (IRB Protocol #: 2018P002864).

